# COVID-19 vaccine confidence and hesitancy among healthcare workers: a cross-sectional survey from a MERS-CoV experienced nation

**DOI:** 10.1101/2020.12.09.20246447

**Authors:** Mazin Barry, Mohamad-Hani Temsah, Abdullah Alhuzaimi, Nurah Alamro, Ayman Al-Eyadhy, Fadi Aljamaan, Basema Saddik, Ali Alhaboob, Fahad Alsohime, Khalid Alhasan, Abdulkarim Alrabiaah, Ali Alaraj, Rabih Halwani, Amr Jamal, Sarah Alsubaie, Fatimah S. Al-Shahrani, Ziad A. Memish, Jaffar A. Al-Tawfiq

**Affiliations:** Division of Infectious Diseases, Department of Internal Medicine, College of Medicine, King Saud University and King Saud University Medical City, Riyadh, Saudi Arabia; Pediatric Department, College of Medicine, King Saud University, Riyadh, Saudi Arabia; Specialty Internal Medicine and Quality Department, Johns Hopkins Aramco Healthcare, Dhahran, Saudi Arabia; Infectious disease division, Department of Medicine, Indiana University School of Medicine, Indiana, USA; Infectious Disease Division, Department of Medicine, Johns Hopkins University School of Medicine, Baltimore, MD, USA; College of Medicine, King Saud University, Riyadh, Saudi Arabia; Division of Pediatric Cardiology, Cardiac Science Department, College of Medicine, King Saud University, Riyadh, Saudi Arabia; Department of Family and Community Medicine, King Saud University Medical City, Riyadh, Saudi Arabia; Critical Care Dept, College of Medicine, King Saud University, Riyadh, Saudi Arabia; Dr. Sulaiman Al Habib Medical Group, Riyadh, Saudi Arabia; College of Medicine, University of Sharjah, Sharjah, UAE; Department of Medicine, College of Medicine, Qassim University, Qassim, Saudi Arabia; Director Research and Innovation Centre, King Saud Medical City, Ministry of Health & College of Medicine, Alfaisal University, Riyadh, Kingdom of Saudi Arabia; Hubert Department of Global Health, Rollins School of Public Health, Emory University, Atlanta, GA, USA

**Keywords:** COVID-19, vaccine, acceptance, confidence, healthcare workers

## Abstract

**Objectives:** This study aimed to identify COVID-19 vaccine perception, acceptance, confidence, hesitancy, and barriers among healthcare workers (HCW).

**Methods:** An online national cross-sectional pilot-validated questionnaire was self-administered by HCW in Saudi Arabia, a nation with MERS-CoV experience. The main outcome variable was HCW’s acceptance of COVID-19 vaccine candidates. The associated factors of vaccination acceptance were identified through a logistic regression analysis and the level of anxiety using generalized anxiety disorder 7.

**Result:** Out of 1512 HCWs who completed the study questionnaire—944 (62.4%) women and 568 (37.6%) men—1058 (70%) were willing to receive COVID-19 vaccines. Logistic regression analysis revealed that male HCWs (ORa=1.551, 95% CI: 1.122–2.144), HCWs who believe in vaccine safety (ORa=2.151; 95% CI:1.708–2.708), HCWs who believe that COVID vaccines are the most likely way to stop the pandemic (ORa=1.539; 95% CI: 1.259–1.881), and HCWs who rely on Centers for Disease Control and Prevention website for COVID 19 updates (ORa=1.505, 95% CI: 1.125–2.013) were significantly associated with reporting willingness to be vaccinated. However, HCWs who believed vaccines were rushed without evidence-informed testing were found to be 60% less inclined to accept COVID-19 vaccines (ORa=0.394, 95% CI: 0.298– 0.522).

**Conclusion:** Most HCWs are willing to receive COVID-19 vaccines once available; yet, satisfactoriness of COVID-19 vaccination among HCWs is crucial because health professionals’ knowledge and confidence toward vaccines are important determining factors for their own vaccine acceptance and recommendation to their patients.

## Introduction

On December 31, 2019, a cluster of pneumonia cases was reported in Wuhan city, Hubei Province, China, and linked to a wet seafood market. Subsequently, a new coronavirus was identified as the etiological agent and named severe acute respiratory syndrome coronavirus-2 (SARS-CoV-2), the causative agent of coronavirus disease 2019 (COVID-19) [1-3]. The World Health Organization (WHO) International Health Regulation Emergency Committee declared COVID-19 a Public Health Emergency of International Concern on January 30, 2020, and a pandemic on March 11, 2020 [4].

As of November 29, 2020, COVID-19 has been reported globally in 191 countries, with 62,311,483 laboratory-confirmed cases causing 1,453,467 deaths [5]. Efforts to eliminate SARS-CoV-2 would be unsuccessful in the long term, as they are constantly challenged by births of new susceptible hosts and waning immunity in previously infected individuals. The durability of SARS-CoV-2 immunity is not yet fully established [6], but births will promote virus survival, thus, similar to other infectious pathogens, SARS-CoV-2 is likely to circulate in humans for many years to come [7].

An unprecedented effort to develop a vaccine started very early in the pandemic to curb the current global situation [8]. Research gaps needed to address the response to COVID-19 have been identified and facilitated work on animal models for vaccine research and development [9]. Different countries and organizations are developing new platform technologies that would support rapid development of such vaccines from viral sequencing to clinical trials in less than 16 weeks, demonstrate elicitation of consistent immune response, and be suitable for large-scale production. Of the greatest potential are DNA- and RNA-based vaccine platforms, which can be developed quickly because they use synthetic processes and do not need cell culture or fermentation, in addition, the use of next-generation sequencing and reverse genetics may also cut the development time of more conventional vaccines [10, 11]. As per WHO, 149 vaccines are currently in preclinical development and 38 candidate vaccines are undergoing evaluation in clinical trials, with multiple vaccines having concluded phase1/2 trials and are already in phase 3 trials. These include JNJ-78436735 an adenovirus vaccine (Ad26.COV2.S)[12][13], mRNA-1273 an mRNA vaccine[14], AZD1222 an adenovirus vaccine (ChAdOx1 nCoV-19)[15], BNT162b1an mRNA vaccine[16], NVX□CoV2373 a full-length recombinant SARS CoV-2 glycoprotein nanoparticle vaccine adjuvanted with Matrix M [17], Ad5-nCoV an adenovirus vaccine [18][19, 20][21]. If one or more of these vaccines are proven to be safe, effective, and approved by regulatory authorities for usage, it would be unlikely to be widely available with sufficient quantities to cover the whole population, hence, a phased approach for vaccine allocation has been developed starting with Phase 1a or “Jumpstart Phase,” targeting high-risk healthcare workers (HCW) and first responders [22, 23].

The Kingdom of Saudi Arabia (KSA) is one of the top thirty countries with the highest reported COVID-19 cases: 356,911 laboratory confirmed cases and 5,870 deaths [5] as of November 29, 2020. It is also one of the few countries in the world in which a second coronavirus, the Middle East respiratory syndrome coronavirus (MERS-CoV) is still causing seasonal epidemics since its discovery in 2012 [24]. As of June 1, 2020, a total of 2,167 laboratory confirmed cases of MERS-CoV were reported in KSA with 842 deaths [25, 26]. An adenovirus-based vaccine against MERS-CoV in dromedary camels was recently developed [27]and is currently in phase 3 trials in KSA. Perception, confidence, and hesitancy for newly developed vaccines in the context of emerging viral infections and pandemics are principal factors in assessing vaccine acceptance. Acceptance of a potential COVID-19 vaccine assessed among the general population of KSA in a survey of 3,101 participants which showed an acceptance rate of 45% among the general public [28], while another public survey among 992 participants revealed an acceptance rate of 65% [29]. However, none of these surveys specifically targeted HCWs who are expected to be included in the jumpstart phase of vaccination. In this study, we investigated COVID-19 vaccine perception, acceptance, confidence, hesitancy, and barriers among HCWs in the KSA.

## Methods

### Data Collection

This was a national cross-sectional survey among HCWs in KSA during the COVID-19 pandemic. [30] Data were collected between 4 and 14 November 2020. Participants were recruited from several social media platforms and email lists using a convenience sampling technique. The survey was a pilot-validated, self-administered questionnaire that was sent to HCW online through SurveyMonkey©, a platform that allows researchers to deploy and analyze surveys via the web [31]. The questionnaire was adapted from our previously published study [26] with modifications and additions related to COVID-19 vaccine candidates. The questions addressed the demographic characteristics of respondents (job category, age, sex, years of clinical experience, and work area), previous exposure to MERS-CoV or COVID-19 infected patients, and whether HCWs themselves were ever infected with COVID-19. We assessed HCW readiness to receive the COVID-19 vaccine as the main outcome. We also evaluated the timing of HCW acceptance to receive the vaccine, HCWs’ beliefs about COVID-19 vaccination, and the barriers and reasons for refusal of new vaccines for those who completely rejected receiving it.

Additionally, we assessed the HCWs’ perceived worry about the COVID19 pandemic using a series of Likert-like scales (Scale 1–5) and their generalized anxiety level using the general anxiety disorder-7 (GAD-7). This validated instrument is a seven-item tool that is used to assess the severity of generalized anxiety disorder, with each item asking the individual to rate the severity of his or her symptoms over the past two weeks [32]. GAD-7 was previously used to assess HCWs’ anxiety due to COVID-19 [33].

Before participation, the purpose of the study was explained in English at the beginning of the online survey. The respondent was given the opportunity to ask questions via a dedicated email address for the study. The Institutional Review Board at the College of Medicine and King Saud University Medical City approved the study (approval # 20/0065/IRB). A waiver for signed consent was obtained since the survey presented no more than a minimal risk to subjects and involved no procedures for which written consent is usually required outside the study context. To maximize confidentiality, personal identifiers were not required.

### Statistical analysis

Descriptive statistics approaches with mean and standard deviation were applied to continuous variables, while percentages were used for dichotomous variables. The two-sample t test was used to evaluate continuous scores, and the chi-squared test (χ^2^) of independence was used to compare proportions.

A multivariate binary logistic regression model was used to explore the associations between the outcome variable of HCWs’ willingness to receive the COVID-19 vaccine and demographic characteristics of HCWs, HCW beliefs toward COVID-19 vaccines and anxiety from COVID-19 and levels of anxiety using the GAD-7. The associations between predictors and the outcome were expressed as adjusted odds ratio and 95% confidence interval. The SPSS IBM Version 21 was [34] used for data analysis, the Excel program was used for creating figures and depictions, and the p-value ≤ 0.050 was considered statistically significant.

## Results

A total of 2,079 HCWs were invited to participate in the study, and 2,007 (96.5%) agreed to participate. Complete data from 1,512 (75.3%) participants were included in the analysis. The details of respondents’ sociodemographic and professional characteristics are depicted in Table 1. Of the respondents, 360 (23.8%) had one or more chronic illnesses, and 194 (12.8%) reported a history of COVID-19 infection confirmed by polymerase chain reaction (PCR). Most (86%) HCWs had been exposed to patients with COVID-19, and almost one third were in contact with family member(s) who had COVID-19 infection. There were 140 HCWs (12.4%) who reported contact with MESR-CoV infected patients as well (Supplementary Table S1).

**Table 1:**
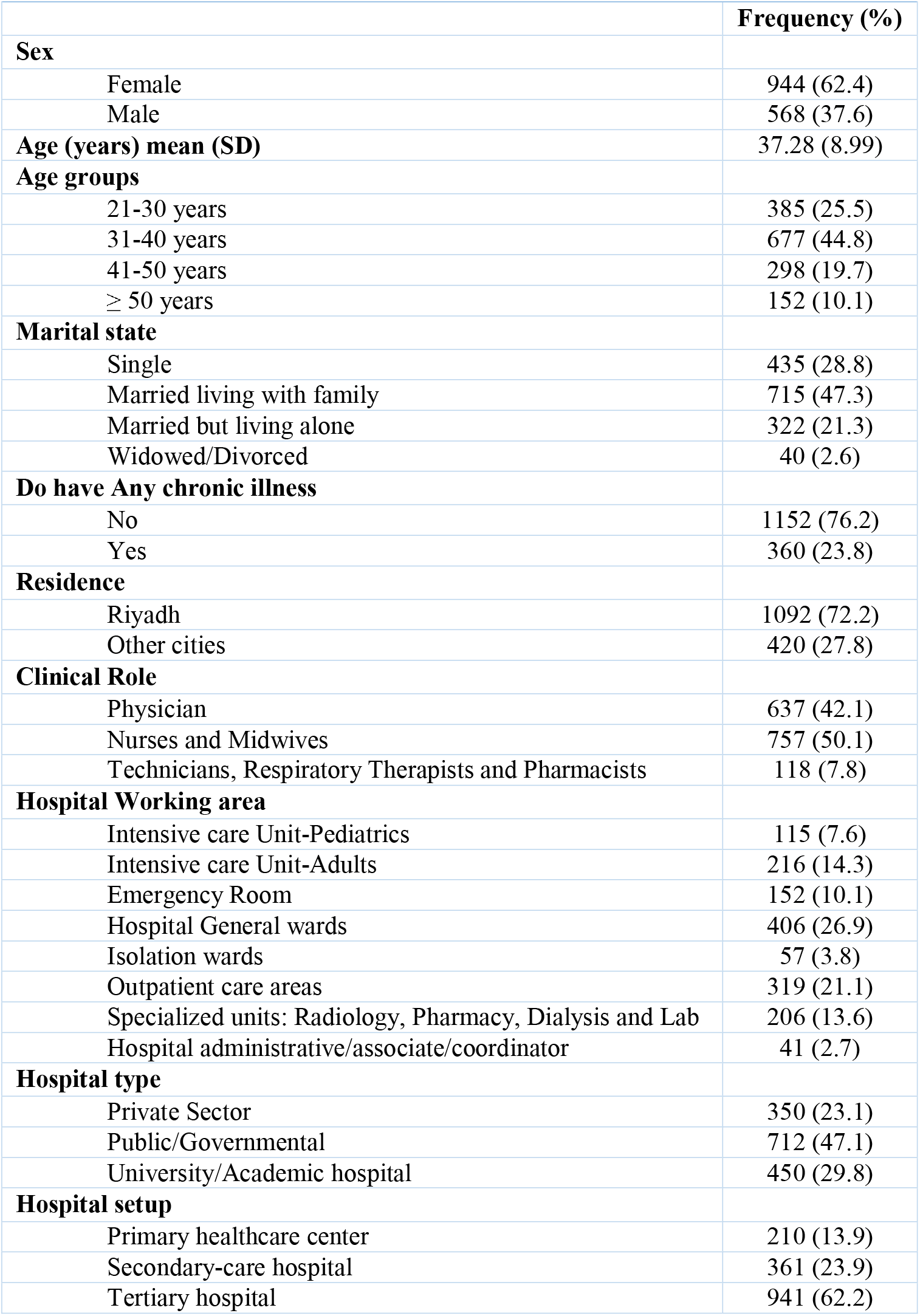
Respondents’ sociodemographic and professional characteristics (N = 1512)

### Acceptance of potential COVID-19 vaccine

Of the 1,512 respondents, 1,058 (70%) were willing to receive a COVID-19 vaccine once available. In terms of readiness to receive the vaccine immediately, most respondents 795 (52.6%) indicated willingness to receive a vaccine as soon as possible, while 35.6% preferred waiting for a few months before receiving it and 11.8% indicated they would never accept receiving any potential vaccine. The majority (83.9%) of respondents reported receiving the annual influenza vaccine over the last two years (Supplementary Table S1). Moreover, 63.7% of HCWs also indicated their willingness to receive a MERS-CoV vaccine if it becomes available. The HCW beliefs about COVID-19 vaccines were evaluated using three statements about the safety of the vaccine, role of vaccine to stop the pandemic, and role of the vaccine in preventing COVID-19 complications. Most HCWs agreed that a vaccine would be safe and would be the best way to stop the pandemic and prevent disease complications. (details are shown in Supplementary Table S2).

Bivariate analysis of participants’ characteristics and their willingness to receive COVID-19 vaccines showed a significant correlation with multiple factors (Table 2): male HCWs (P = 0.0022) and being married but living alone (P = 0.016) were significantly associated with willingness to receive a COVID-19 vaccine. Those who were willing to receive the vaccine had significantly higher general anxiety scores and specific anxiety from contracting COVID-19 infection or transmitting it to their family members (Table 2). These HCWs significantly agreed that once a vaccine is available, it would be safe and thought it would be the best way to stop the pandemic and avoid disease complications.

**Table 2:**
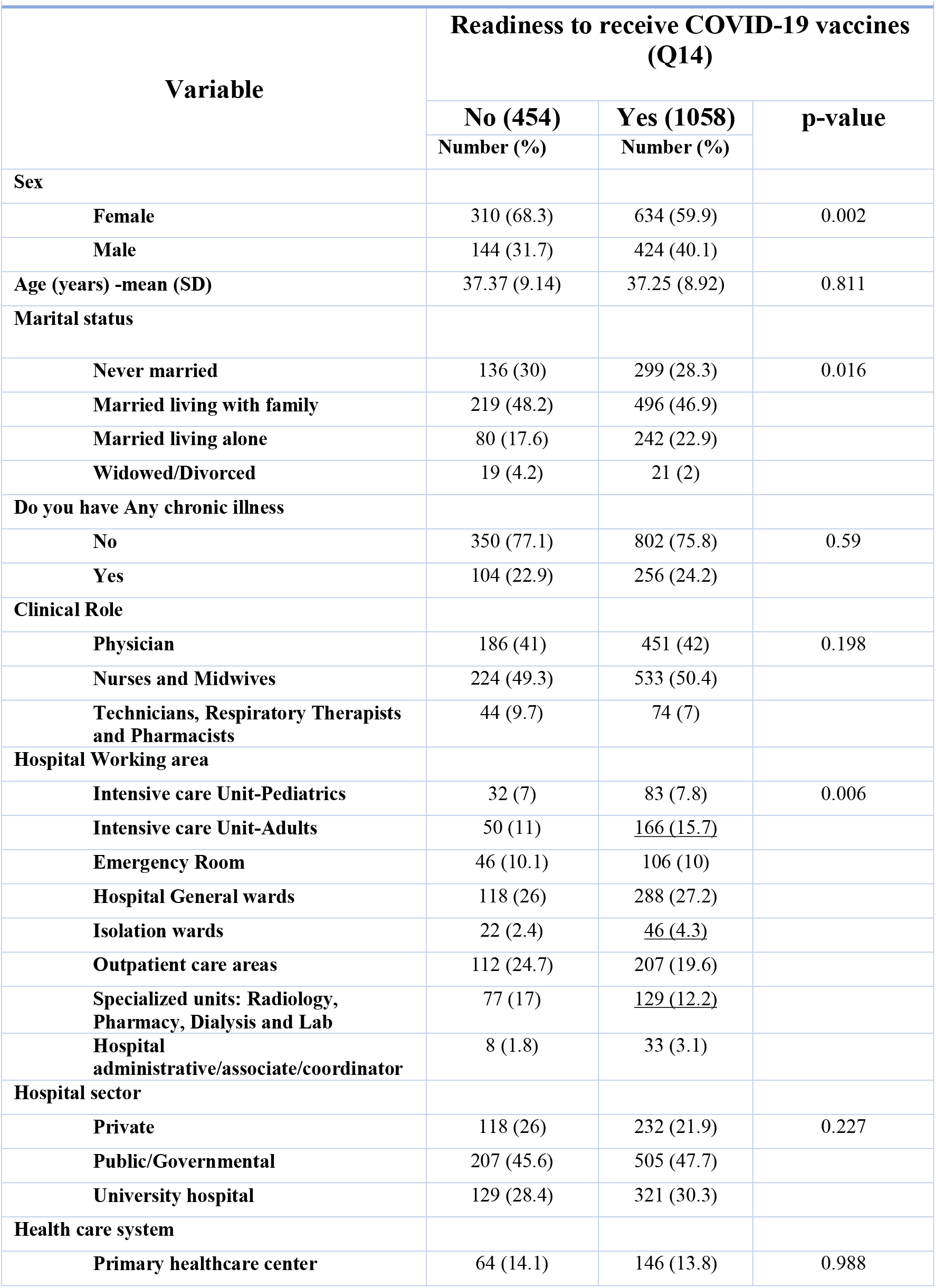
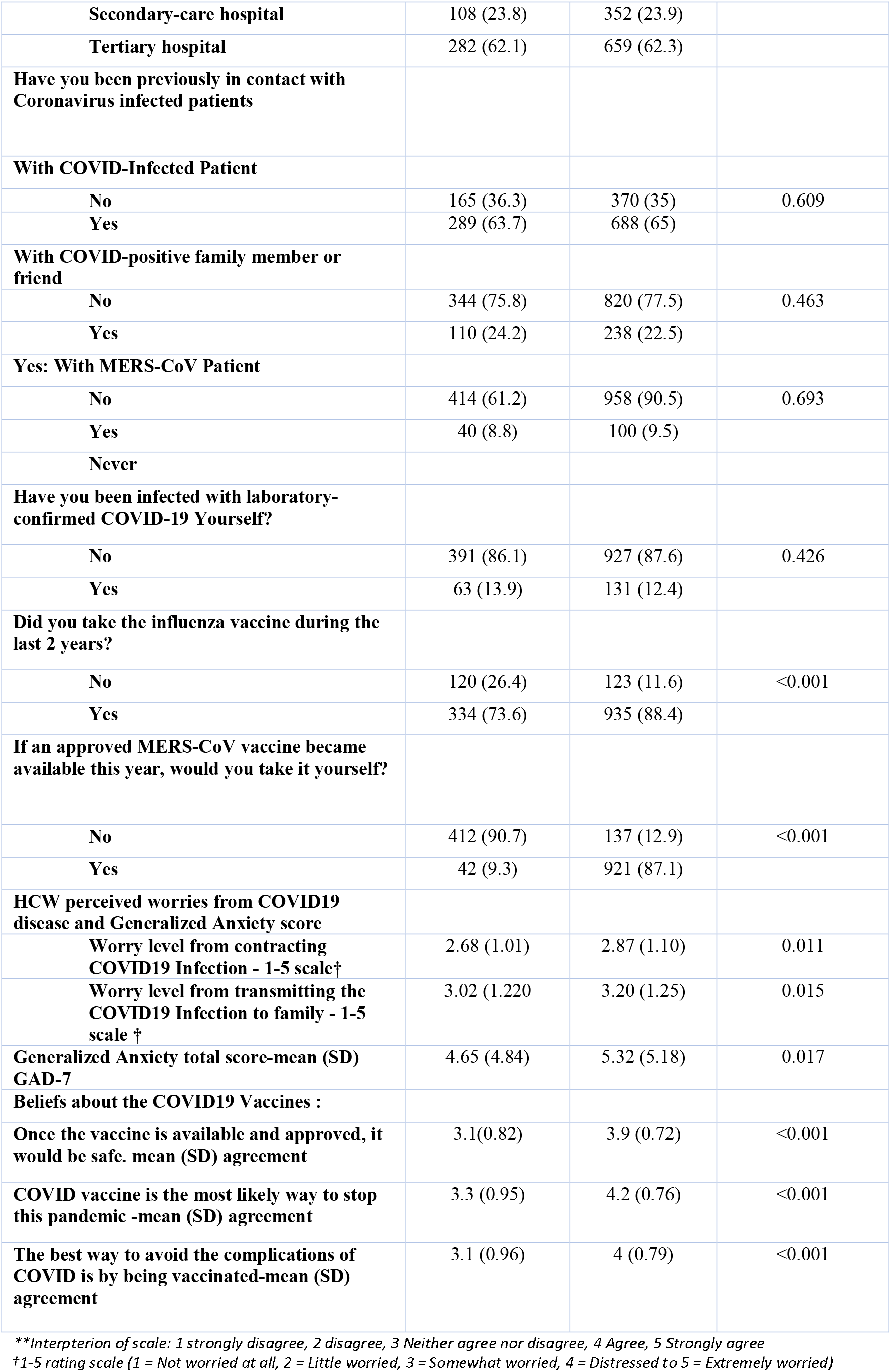
Bivariate analysis of healthcare workers’ willingness to receive potential COVID-19 vaccines (N = 1512).

HCWs working in adult intensive care units and isolation floors were significantly associated with higher readiness to receive the vaccine (p = 0.006). HCWs who took their annual influenza vaccine in the last two years were significantly more likely to accept COVID-19 vaccines. Further, HCWs who were willing to receive a MERS-CoV vaccine once available were also likely to accept the potential COVID-19 vaccine.

### Multivariate analysis of the HCW’s willingness to receive a COVID-19 vaccine

The predictors of HWC’s willingness to accept a COVID-19 vaccine were analyzed using a multivariate binary logistic regression. Table 3 shows the adjusted odds ratio for the various characteristics. Males were 1.55 times more likely to accept a COVID-19 vaccine than females (p = 0.008). Other sociodemographic characteristics, such as age, marital status, previous personal COVID-19 infection, and presence of chronic medical illness, as well as their clinical role and clinical working area, did not correlate significantly with willingness to receive a COVID-19 vaccine.

**Table 3:** Multivariate binary logistic regression analysis of the HCWs’ willingness to get COVID19 vaccine(s) (N=1512)

|  | Multivariate Adjusted Odds Ratio | 95% C.I. for O.R |  | p-value |
| --- | --- | --- | --- | --- |
|  |  | Lower | Upper |  |
| Sex= Male | 1.551 | 1.122 | 2.144 | .008 |
| Age (years) | 1.004 | .987 | 1.021 | .654 |
| Marital state= Never married | 1.084 | .803 | 1.463 | .600 |
| Previously infected with COVID19 disease | .897 | .605 | 1.330 | .588 |
| Chronic medical disease | 1.195 | .860 | 1.660 | .289 |
| Clinical Role | .855 | .671 | 1.091 | .208 |
| Hospital working area | .958 | .894 | 1.028 | .233 |
| Belief in vaccine safety once approved and released (score 1-5) | 2.151 | 1.708 | 2.708 | <0.001 |
| Belief that COVID vaccine is the most likely way to stop the pandemic (score 1-5) | 1.539 | 1.259 | 1.881 | <0.001 |
| Belief that COVID-19 vaccination would be the best way to prevent disease complications (score 1-5) | 1.484 | 1.208 | 1.823 | <0.001 |
| Belief that vaccines are being rushed without testing (score 1-5) | .394 | .298 | .522 | <0.001 |
| Worry level from contracting COVID19 Infection -mean score | 1.170 | .988 | 1.387 | .069 |
| Worry level from transmitting COVID19 Infection to family - mean score | 1.049 | .909 | 1.210 | .512 |
| Generalized Anxiety Score (GAD-7) mean score | 1.016 | .986 | 1.047 | .306 |
| Working in a University Hospital | 1.133 | .842 | 1.525 | .410 |
| Relies on CDC website for information on COVID19 disease updates. | 1.505 | 1.125 | 2.013 | .006 |
| Constant | .006 |  |  | <0.001 |
| *Dependent variable= (Willingness to take COVID19 vaccine No/Yes)* |  |  |  |  |

The participants’ belief in a vaccine’s safety once approved by regulatory authorities correlated significantly with their readiness to accept a vaccine (2.15 times; p<0.001). Likewise, their belief that a vaccine could stop the COVID-19 pandemic and prevent disease complications correlated significantly with their willingness to accept a vaccine (1.5 times, p<0.001). By contrast, HCWs who believed that vaccine candidates were rushed without evidence-informed testing were found to be 60% less inclined to accept a COVID-19 vaccine once available (p<0.001).

Levels of worry about being infected with or transmitting the disease and general anxiety level did not correlate significantly with willingness to accept a COVID-19 vaccine in multivariate analysis. Lastly, HCWs who used the Centers for Disease Control and Prevention (CDC) website to seek evidence-informed knowledge on COVID-19 vaccines were 1.51 times more likely to accept potential vaccine candidates than HCWs who used other sources of information (p = 0.006).

### Reasons for unwillingness to receive a Covid19 vaccine

Almost 12% of HCWs reported they would never agree to receive any COVID19 vaccine candidate (n=177). When asked for reasons for refusal, the most implicated were inadequate data on the safety of a new vaccine and concerns of adverse effects (Table 4).

**Table 4:** Reasons for unwillingness to receive COVID19 vaccines (N = 177)

| Reasons | Number (%) |
| --- | --- |
| Inadequate data about the safety of a new vaccine | 127 (71.8) |
| A concern on adverse effects of the vaccine | 87 (49.2) |
| A concern on vaccine being ineffective | 35 (19.8) |
| Prior adverse reaction to any vaccine | 28 (15.8) |
| I am against vaccines in general | 24 (13.6) |
| **A concern of acquiring Covid19 infection from the vaccine itself** | 24 (13.6) |
| **I perceive myself not to be at considerable risk of developing complications if I am infected with Covid19** | 21 (11.9) |
| **I perceive myself not at elevated risk to acquire Covid19 infection** | 15 (8.5) |
| **I already had COVID infection** | 13 (7.3) |
| **Vaccine administration is painful or inconvenient** | 1 (0.6) |

## Discussion

Understanding the dynamics of vaccine confidence has always been important for public health [35] and is now vital in facing the global COVID-19 pandemic. As many vaccines are currently in phase 3 clinical trials, studying the dynamics of vaccine acceptance is important for planning vaccinations in targeted populations, including HCWs, who would be first to receive the vaccines once approved by regulatory authorities and the ones to advocate and prescribe them to their patients.

Vaccine hesitancy has been documented as a threat to reducing the burden of infectious diseases and has been a cause of resurgence of vaccine preventable diseases. In the context of COVID-19, vaccine hesitancy may cause delays in the acceptance or refusal of new vaccines. In this study, two-thirds of HCWs expressed willingness to receive a potential COVID-19 vaccine. Cited concerns for hesitancy were lack of sufficient safety and efficacy data. Other concerns included potential adverse effects and the belief that a vaccine would be ineffective. Hesitancy is influenced by factors such as complacency and confidence, which affect vaccine acceptance or refusal [36]. We found that HCWs expressed their confidence in a vaccine, as 72% thought that a vaccine would be the most effective way to end the pandemic and 57% thought a vaccine this soon was a scientific achievement. Complacency relates to the perceived risk of disease, as 20% of HCWs who would not receive a vaccine indicated they did not perceive themselves at risk of developing COVID-19 or its complications.

Thirty percent of HCWs were not willing to receive any potential COVID-19 vaccine candidate. These results are consistent with previous research on vaccine confidence for the measles mumps and rubella vaccine, in which only 64% of general practitioners believed the vaccine was safe, while 19% did not believe it was important for children. [3]. Similarly, a cross-sectional survey conducted during the 2009 influenza A (H1N1) pandemic showed an extremely low vaccination rate of 12.7% among HCWs, with most believing it to be unsafe and ineffective [37]. Another survey among 1,340 HCWs revealed that only 58% were willing to recommend the influenza vaccine to their diabetic patients [38]. Developing vaccine confidence among HCWs is a major step in stopping the pandemic amid all the misinformation on different media platforms. Misinformation and distrust are not new for vaccines; conspiracy theories and suspicions about vaccines are common throughout many countries over several decades [36, 37]. A coherent, flexible strategy for COVID-19 vaccination will require unique and collective ingenuity in addressing the public health and immunization needs of HCWs and their patients [41]. Among the 30% of HCWs who were hesitant to receive a COVID-19 vaccine, the top reasons for refusal included the novelty and rapid development of the vaccines and fear of adverse effects, all of which are key questions to be addressed. [42].

Of all the HCWs respondents, 83.9% had received an influenza vaccine, and 63.7% agreed to receive a MERS-CoV vaccine should it be approved. The overall acceptance rate of 70% to a COVID-19 vaccine in our current study is close to a previous report of 64.7% acceptance among all surveyed individuals in KSA [29]. A study from the Republic of Congo showed that only 27.7% of HCWs would accept a COVID-19 vaccine once available [40]. The similarity in accepting a COVID-19 vaccine between HCWs and the general population was previously reported in a study from China, with acceptance rates of 76.4% for HCWs and 72.5% in the general population [44]. These similarities in acceptance rates are interesting and hint that acceptance may not be influenced by professions. As HCWs are expected to receive any approved vaccine first, it is clear from these studies that further education is needed to convey the importance of vaccination and to build confidence to help elevate the acceptance rates among HCWs. One important finding in our study is that 12.8% of the HCW surveyed had been infected with COVID-19. In the KSA, there are no published data on the number or percentage of HCWs among all COVID-19 patients. However, a recent serosurvey showed that the overall seroprevalence rate was 2.36%. [45] In a systematic review, the overall seroprevalence rate of COVID-19 among HCWs was about 11% [43].

In a multivariate logistic regression analysis, males were more likely to accept COVID-19 compared to females (1.55 times higher), although age, marital state, comorbidity, and clinical discipline did not converge significantly on their readiness to receive a COVID19 vaccine. However, in the mentioned study from China, there was no difference in the acceptance rates between males and females [44]. Another study showed that males were more likely to accept vaccination (ORa=1.17) [40]. These differences might be related to the heterogeneity of the populations included in the different studies.

An interesting observation in this study is that HCWs’ belief in the ability of the vaccine to reduce COVID-19 complications predicted significantly greater odds of accepting a vaccine. However, the HCW’s mean worry level from contracting COVID-19 disease and infecting household members did not converge significantly on their odds of willingness to be vaccinated. Since the confidence and hesitancy of HCWs to vaccines are crucial factors in their likelihood of being advocates to vaccinating their patients, this and other similar studies highlight the need for more education and improvement in vaccine confidence among HCW [42].

### Limitations

There are several limitations to this study. First, it was done using convenience sampling; therefore, it cannot be generalized to the entire population. However, we believe that national outreach to recruit HCWs from all regions provides a basis for further national representative studies. Second, this study is subject to the limitations of cross-sectional surveys, including sampling, response, and recall biases. Lastly, the study was done during a period of heavy media coverage on potential COVID-19 vaccines, which could influence the levels of knowledge, perceptions, and attitudes. Despite these limitations, the study highlighted the importance of addressing HCWs’ perceptions and attitudes toward potential COVID-19 vaccines and ensuring the provision of information from trustable sources, which will contribute to better vaccine acceptance among HCWs.

## Conclusion

High acceptance of the COVID-19 vaccine has been shown among HCWs. Concerns about vaccine safety, efficacy, and adverse effects provide important targets for possible interventional educational programs to enhance vaccination rates. Public health authorities and medical organizations need to address this principal issue for a successful vaccination campaign. It is critical that clinicians stay well informed about emerging data on vaccine candidates so that they can help patients make correct decisions about vaccines that are urgently needed to help end the pandemic.

## Data Availability

All the data for this study will be made available upon reasonable request to the corresponding author.

## Supplementary materials

**Supplementary Table S1:**
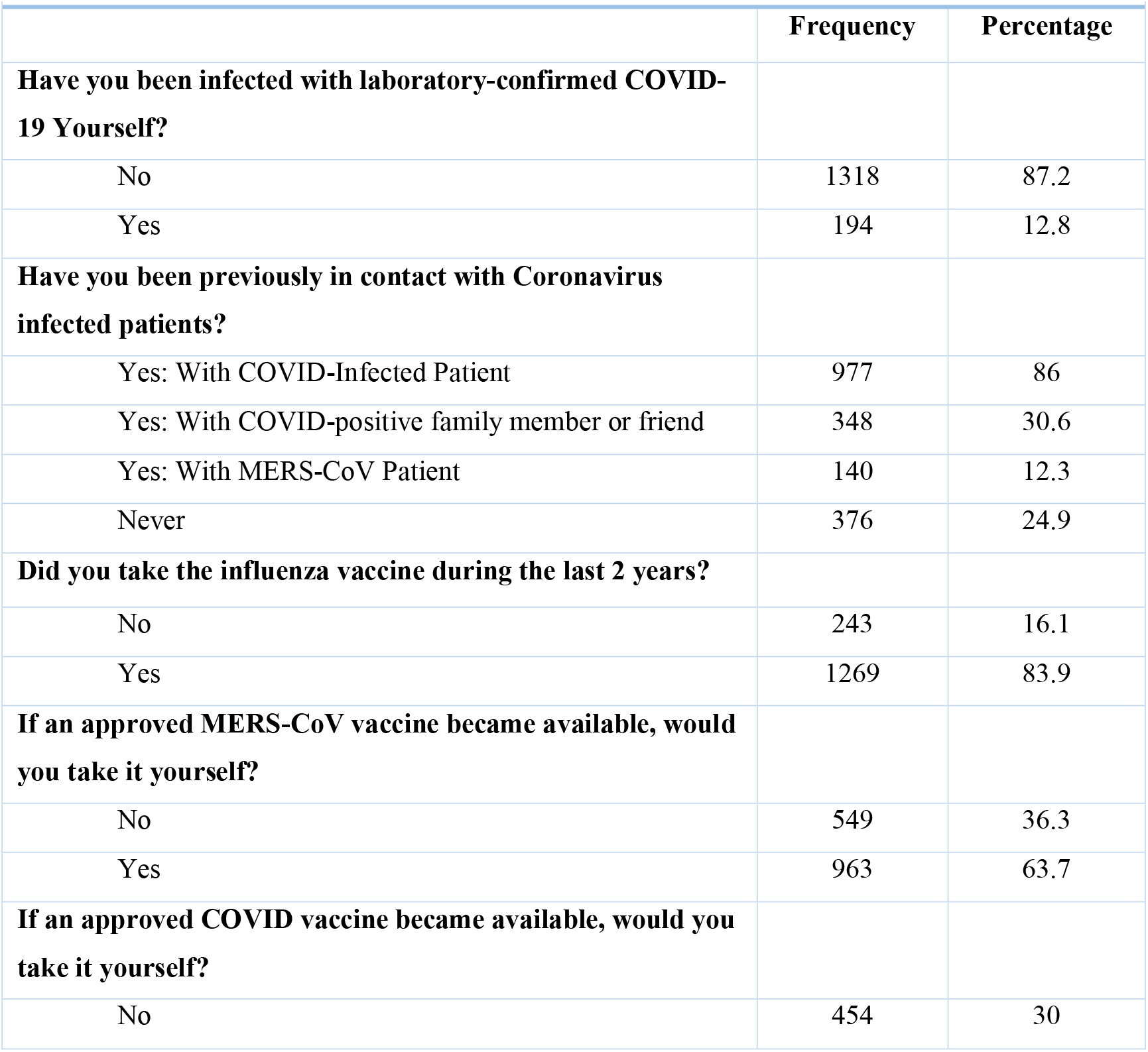
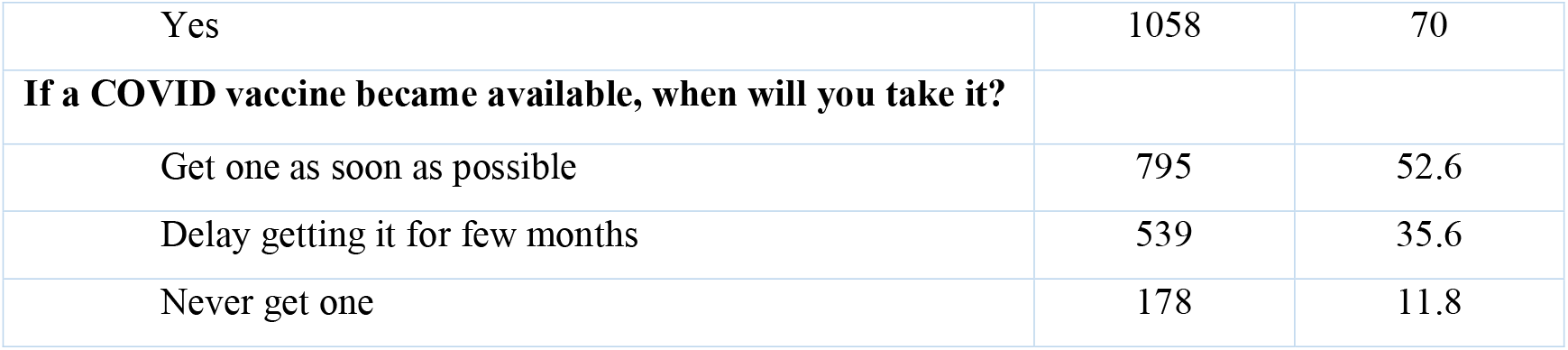
Respondents Attitudes toward COVID-19 vaccine and their experience with COVID-19 pandemic. N = 1512

|  | Frequency | Percentage |
| --- | --- | --- |
| **Have you been infected with laboratory-confirmed COVID-19 Yourself?** |  |  |
| No | 1318 | 87.2 |
| Yes | 194 | 12.8 |
| **Have you been previously in contact with Coronavirus infected patients?** |  |  |
| Yes: With COVID-Infected Patient | 977 | 86 |
| Yes: With COVID-positive family member or friend | 348 | 30.6 |
| Yes: With MERS-CoV Patient | 140 | 12.3 |
| Never | 376 | 24.9 |
| **Did you take the influenza vaccine during the last 2 years?** |  |  |
| No | 243 | 16.1 |
| Yes | 1269 | 83.9 |
| **If an approved MERS-CoV vaccine became available, would you take it yourself?** |  |  |
| No | 549 | 36.3 |
| Yes | 963 | 63.7 |
| **If an approved COVID vaccine became available, would you take it yourself?** |  |  |
| No | 454 | 30 |
| Yes | 1058 | 70 |
| **If a COVID vaccine became available, when will you take it?** |  |  |
| Get one as soon as possible | 795 | 52.6 |
| Delay getting it for few months | 539 | 35.6 |
| Never get one | 178 | 11.8 |

**Supplementary Table S2:** Healthcare workers perceptions/opinion about future COVID-19 vaccines

|  | N (%) |  |  |  |  |
| --- | --- | --- | --- | --- | --- |
| ☐ | Strongly Disagree | Disagree | Undecided | Agree | Strongly Agree |
| Once the vaccine is available and approved, it will be safe. | 20 (1.3) | 81 (5.4) | 544 (36) | 650 (43) | 217 (14.4) |
| COVID-19 vaccine is the most likely way to stop the pandemic. | 17 (1.1) | 75 (5) | 356 (23.5) | 627 (41.5) | 437 (28.9) |
| The best way to avoid complications of COVID-19 is by being vaccinated | 25 (1.7) | 135 (8.9) | 389 (25.7) | 670 (44.3) | 293 (19.4) |

